# Adverse events affecting recovery from seasonal influenza vaccination in the hypertensive population: A population-based pharmacovigilance analysis

**DOI:** 10.1101/2024.09.03.24313030

**Authors:** Hao Wu, Xiaona He, Yu Cao, Wei Gao

## Abstract

Seasonal influenza vaccination is crucial for preventing influenza and its complications. Data from the U.S. Vaccine Adverse Event Reporting System (VAERS) indicate a higher proportion of adverse events (AEs) after influenza vaccination in hypertensive people. However, there is limited evidence on AEs in hypertensive people following seasonal influenza vaccination. We collected a total of 4647 individuals on seasonal influenza vaccination and 6380 AEs from VAERS for hypertensive people aged 18 years or older from 1 January 2013 to 23 June 2023. We conducted propensity score matching (PSM) by adjusting for the demographic characteristics of the seasonal influenza-vaccinated hypertensive population and the season of onset. Cox regression analysis was used to calculate the risk ratio of reported adverse events (AEs) that affected recovery after seasonal influenza vaccination. Most AEs were nonserious and occurred within 48 hours. The most common AEs were general disorders and administration site conditions (therapeutic and non-therapeutic responses, inflammation) and musculoskeletal and connective tissue disorders (musculoskeletal and connective tissue pain and discomfort, bursal disorders, joint-related signs, and symptoms). All three types of seasonal influenza vaccines were associated with injection site reactions (47.07% trivalent influenza vaccine [TIA], hazard ratio, HR 2.04, 95% confidence interval, CI 1.22–3.40; 20.00% quadrivalent influenza vaccine [QIA], HR 2.81, 95% CI, 1.81–4.37; 67.48% influenza vaccine, unknown manufacturer [FLUX], HR 2.83, 95% CI, 1.12–7.15) and were the AEs affecting the largest proportion of delayed recoveries in the hypertensive population. Potential AEs following seasonal influenza vaccination may affect the recovery of the hypertensive population. The majority of AEs reported were general disorders, predominantly injection site reactions, and nonserious.

## Introduction

Influenza is a highly transmissible viral infection caused by the influenza virus, leading to severe respiratory illnesses in approximately 3–5 million people and 290,000– 650,000 deaths globally every year[1,2]. Influenza has become a serious public health issue. Since the 20th century, several influenza pandemics have occurred in human history, such as the ‘Spanish Flu’ in 1918–1919, the ‘Asian Flu’ in 1957–1958, the ‘Hong Kong Flu’ in 1968–1969, the ‘Swine Flu’ in 1976, the ‘Russian Flu’ in 1977, and the Influenza A in 2009, which also caused millions of deaths[3,4]. Pandemic and seasonal influenza cause substantial morbidity and mortality worldwide, posing a serious threat to public health and the global economy[5]. Seasonal influenza infections occur in all age groups, but children under 59 months of age, adults over 65 years of age, and pregnant women are more vulnerable[6–8]. Despite the high incidence of annual seasonal influenza infections, annual vaccination programs remain a critical public health tool to reduce the disease burden of seasonal influenza[9]. Currently, seasonal influenza vaccines are produced as ‘trivalent’ or ‘quadrivalent’ formulations[10,11]. Seasonal influenza vaccines are generally considered safe; however, they can sometimes cause adverse events following immunization (AEFIs). Generally, AEFI is deemed less serious than influenza[12]. Therefore, pharmacovigilance (PV) is an important tool for monitoring AEFI and confirming the benefits of immunization in different target groups[13].

The U.S. Vaccine Adverse Event Reporting System (VAERS) was created in 1990 and co-administered by the U.S. Centers for Disease Control and Prevention (CDC) and the U.S. Food and Drug Administration (FDA) to collect spontaneous reports of adverse events (AEs) and manage vaccine safety VAERS accepts reports of AEs from health-care providers, vaccine manufacturers, and the public[14,15]. The main objectives of VAERS are to detect new, unusual, or rare vaccine AEs; assess the safety of newly approved vaccines; make new recommendations for existing vaccines; identify potential risk factors, monitor the increase in known AEs; and determine and address possible reporting clusters[16]. Real-world monitoring of vaccine safety is crucial for understanding suspected AEFIs, assessing their incidence, and determining risk factors to identify potential vaccine contraindications[17]. Moreover, the recently published World Health Organization (WHO) guidelines on the economic evaluation of influenza vaccination suggest considering AEFI when possible[18].**Error! Reference source not found.**

Although influenza is mostly self-limiting, serious complications can occur in vulnerable patient groups[12]. Hypertension is an established cardiovascular risk factor for the general population. Patients with hypertension are regarded as a high-risk group for being infected with seasonal influenza due to the high possibility of a compromised immune system, especially in the elderly[19,20]. The WHO[1] and U.S. CDC[21] recommend that patients with chronic diseases should be priority groups for influenza vaccination. The Chinese CDC has recommended that it is of vital importance for older hypertensive patients to receive an influenza vaccine[22], which is consistent with the recommendations of the U.S. CDC[23]and the WHO. One study showed that the coverage rate of influenza vaccinations in which only 0.4% of people with chronic diseases had received an influenza vaccine[24], a history of influenza vaccination, and the perceived safety and effectiveness of the vaccination are responsible for the lower values of influenza vaccination coverage in mainland China[25,26].

Thus, the objective of our study was to better understand the AEs affecting recovery from seasonal influenza vaccination in the hypertensive population. We investigated all possible AEs for the three types of seasonal influenza vaccines (trivalent influenza vaccine [TIA], quadrivalent influenza vaccine [QIA], and influenza vaccine, unknown manufacturer [FLUX]) between January 1, 2013, and June 23, 2023 on the VAERS database.

## Materials and Methods

### Study data selection

A total of 81,713 AEs were initially investigated during the study period; of these, 4647 individuals with a history of hypertension with a unique VAERS ID were screened: 2135 for TIA, 2300 for QIA, and 212 for FLUX. The primary endpoint was all AEs within one month in individuals with a history of hypertension. The exclusion criteria were missing data on age or being under 18 years of age, missing data on the number of days associated with vaccine-related AEs, cases reported after one month, failure to report recovery from vaccine-induced AEs, outliers or flawed logic, and data for which the year of vaccination did not coincide with the year reported. S1 Fig displays the workflow, illustrating the data collection process and the number of individuals excluded at different stages.

### Study population

Data from the original reports are publicly available as an online downloadable dataset on the VAERS website, with sensitive patient information removed[14]. The raw dataset comprised three distinct files in the VAERS database between January 1, 2013, and June 23, 2023. It included demographic information and medical history in VAERSDATA, vaccination-associated adverse symptoms in VAERSSYMPTOMS, and vaccine type in VAERSVAX. We identified all cases related to the seasonal influenza vaccine by using a unique case identification number in the raw dataset. We examined the medical records for each individual’s medical history, particularly focusing on pre-diagnosed diseases at the time of seasonal vaccination. We used keywords such as high blood pressure, hypertension, HBP, and high blood pressure to extract individuals with a history of high blood pressure as the primary population. The analysis of medical records included general information (VAERS ID, age, sex, state, and medical history when vaccinated), vaccination information (vaccine type, vaccination date, AEs onset date, and the interval between the vaccination date and the occurrence of AEs), and recovery from vaccine-induced AEs.

### Selection of AE reports

According to the unique ID identification number, 4647 individuals with a history of hypertension were included after the deletion of blank values, age under 18 years, and certain records with logical errors. Additionally, we examined five adverse symptoms for each participant to more precisely measure the AEs affecting their recovery from seasonal influenza vaccination. Seasonal influenza vaccine AEs were defined as reports submitted that contained the word ‘FLU’, but did not specifically mention H1N1 influenza in the vaccine code. To compare the different types of seasonal influenza vaccines, we classified them into three primary categories: (1) TIA, including FLU3, FLUA3, FLUC3, FLUR3, and FLUN3; (2) QIA, including FLU4, FLUA4, FLUC4, FLUR4, and FLUN4; and (3) FLUX with no brand name.

### Standard medical terms for vaccine AEs

The Medical Dictionary for Regulatory Activities (MedDRA) is a medical coding dictionary used by regulatory authorities, pharmaceutical companies, clinical research organizations, and health care professionals[27]. MedDRA is organized into a five-level hierarchy, with the highest level being the Systematic Organ Classification (SOC), which is further divided into High-Level Group Terms (HLGT), High-Level Terms (HLT), Preferred Terms (PT), and Low-Level Terms (LLT). In release version 26.1[28], the hierarchy consists of 27,337, 1738, 24,313, and 81,885 terms. The VAERS database provides several AEs at the PT level, but these are highly fragmented into signs, symptoms, diagnoses, investigations, or medical procedures, which may fail to recognize differences in the incidence of AEs[29]. Therefore, we converted the standard terminology associated with all AEs (PT) into HLT[30]. AEs at the primary SOC level were selected for MedDRA’s hierarchical structure analysis.

### Statistical analysis

The VAERS database collected data on age groups, sex, region, and season of onset. The type of vaccine used may depend on individual characteristics; therefore, individual choices may influence vaccine-associated AEs and recovery. Therefore, we applied propensity score matching (PSM) to reduce bias due to these confounding variables. A nearest-neighbor approach with a 1:1 matching ratio was used. PSM shifts the regulation of confounders to the control of propensity scores to ‘downscale’ and to control for confounding bias. The propensity score (PS) for seasonal influenza vaccine exposure was estimated using logistic regression modeling with age group (18-59, 60-64, 65-69, 70-74, 75-79, and ≥ 80 years), region (West, Midwest, Northeast, South, and Overseas territory), sex (female and male), and season of onset (spring including March, April, and May; summer including June, July, and August; fall including September, October, and November; and winter including December, January, and February) as predictors. To control for confounding variables, estimates were obtained using a binomial logistic regression analysis. The caliper value was defined as ‘0.2×standard deviation of the PS estimates with logit transformation applied’.

Standardized mean difference (SMD) was calculated to assess variable balance in the matched data, and a SMD value <0.1 was considered balanced[31]. Using PS estimates and caliper values, the hypertensive population seasonal influenza vaccine recovery and non-recovery groups were matched. The case selection for proportional imbalance analysis is shown in S1 Fig. A total of 6380 seasonal influenza vaccine-induced AEs reported for each individual over one month were considered independent variables in our study. The dependent variable was whether the hypertensive population recovered from influenza vaccine-induced AEs or not. Cox proportional hazards regression analysis was used to examine the association between these AEs and recovery. AEs with a total frequency of less than 10 were removed to address any monotonic likelihood issues that may have occurred in the Cox proportional hazards regression models[30]. The results of the Cox regression models are expressed as hazard ratios (HR) and 95% confidence intervals (CI). A consistency index was used to determine the discriminatory power of the multivariate Cox proportional hazard regression model. All statistical analyses were performed using R version 4.3.2 (R Foundation for Statistical Computing, Vienna, Austria). Prism version 9 (GraphPad Software, USA) was used as a mapping tool, and the MedDRA desktop viewer was used for hierarchical structure analysis of the medical terms[29,30].

## Results

Of the 4647 validated cases with a history of hypertension, the mean baseline age of the study participants was 64.46 years. The gender distribution was 3132 females (67.40%), and 1500 males (32.28%), with a male-to-female ratio of approximately 1:2.08. The southern region had the highest number of seasonally influenza-vaccinated hypertensive individuals (34.02%). In contrast, there was little difference in the number of individuals vaccinated in the western (22.04%) and midwestern regions (23.28%). The AE period coincided with the peak season for seasonal influenza vaccination in autumn, which had the highest number of episodes among the four seasons.

Additionally, the AEs episode interval for the seasonal influenza vaccine was 3.30 days: 3.03 for TIA; 3.38 for QIA; and 5.06 for FLUX. The results showed that 2188 (47.08%) patients in the hypertensive population did not recover from AEs (Table 1). After PSM was paired at 1:1 ratio to eliminate selection bias, the caliper width was adjusted to 0.2, resulting in the disappearance of differences between groups (SMD<0.1, P>0.05). The total number of seasonal influenza vaccinations after matching was 3914. We found differences in age, sex, region, and season of onset between the non-recovered and recovered groups in the hypertensive population before PSM for TIA, QIA, and FLUX (S1-S3 Tables).

**Table 1.**
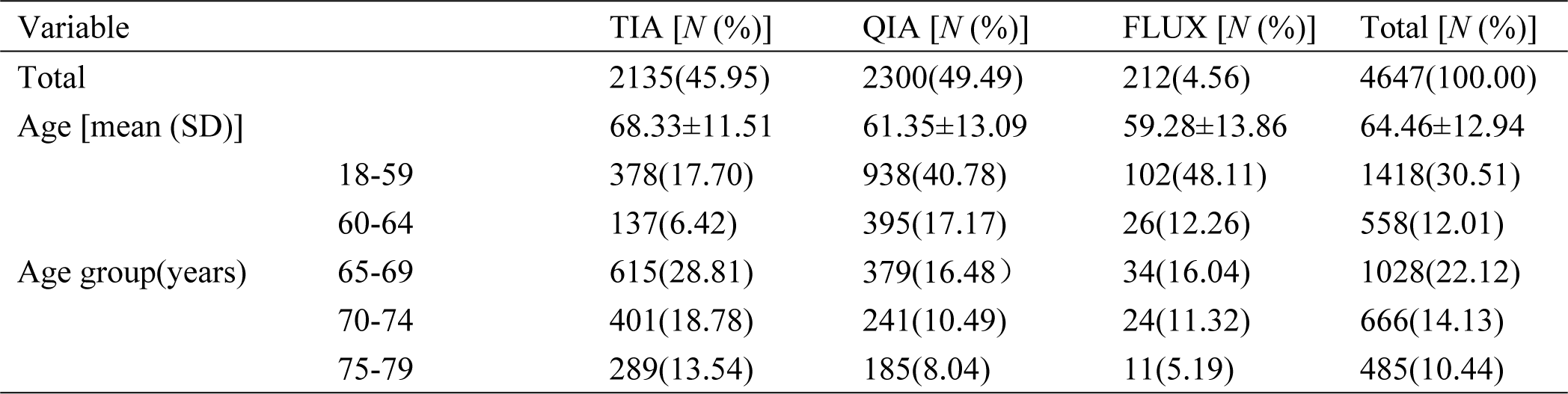
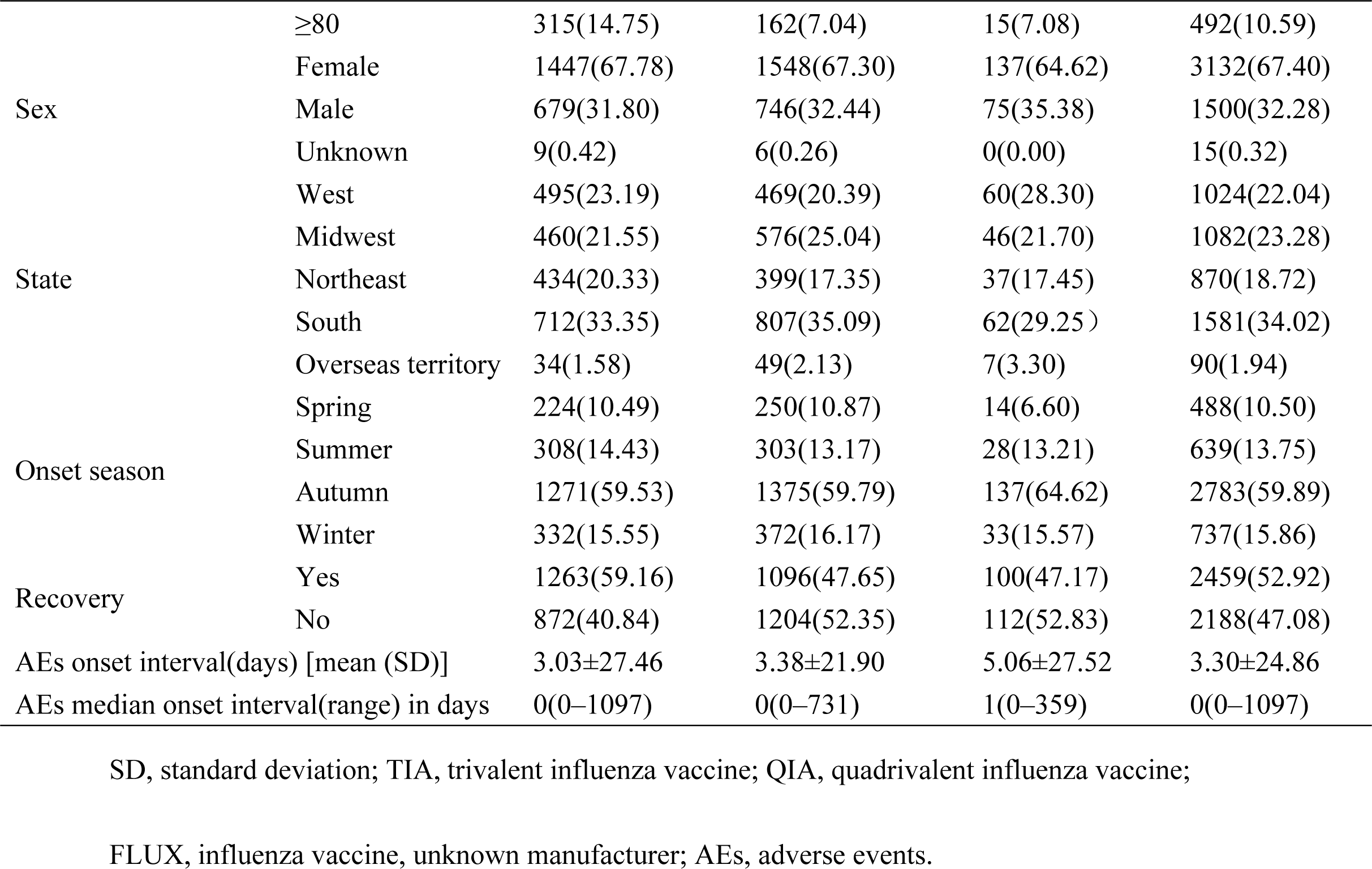
Demographic characteristics of seasonal influenza vaccination in the hypertensive population.

In the PS-matched hypertensive population, 75 AEs with TIA have been reported in the PS-matched hypertensive population. Furthermore, we identified 12 AEs affecting recovery that were associated with the estimated risk of occurrence using a multivariate Cox regression model with a consistency index of 0.58. Overall, 12 AEs were linked to TIA (Fig 1), with an adjusted HR range of 1.82 to 3.91, and were categorized into eight SOC terms (S4 Table). Among the SOC terms, two common AEs accounted for the majority: general disorders and administration site conditions (61.68%) and musculoskeletal and connective tissue disorders (23.85%). Especially, injection site reactions (47.07%; HR=2.04; 95% CI, 1.22–3.40), musculoskeletal and connective tissue pain and discomfort (15.71%; HR=2.40; 95% CI, 1.43–4.10), pain and discomfort NEC (13.11%; HR=1.99; 95% CI, 1.17–3.40) showed an increased risk of non-recovery in hypertensive individuals. After the QIA, 94 AEs were reported, and 57 AEs were estimated using the multivariate Cox regression model, with a consistency index of 0.63 (Fig 2). The adjusted HR distribution ranged from 2.08 to 5.85 and was grouped into 15 SOC terms (S5 Table).

**Fig 1.**
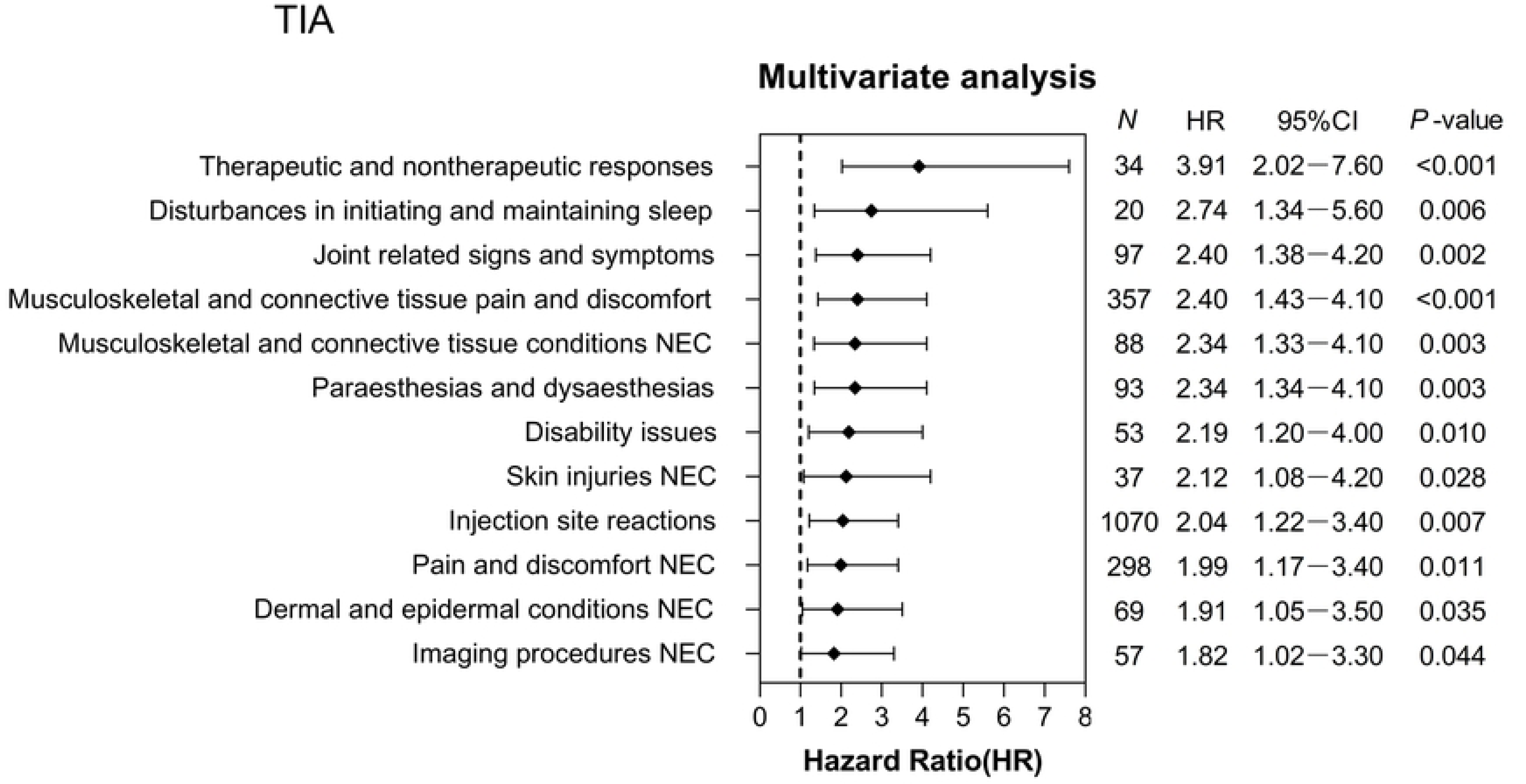
Adverse events (AEs) hazard ratios after trivalent influenza vaccines (TIA) vaccination estimated from a multivariate Cox proportional hazard model. The results showed 12 trivalent influenza vaccine (TIA) vaccine-induced adverse events (AEs) with a statistically significant difference (*P*<0.05), estimated using multivariate Cox proportional hazard regression analysis. N denotes the number of adverse cases in the non-recovered group; HR, hazard ratio; CI, confidence interval.

**Fig 2.**
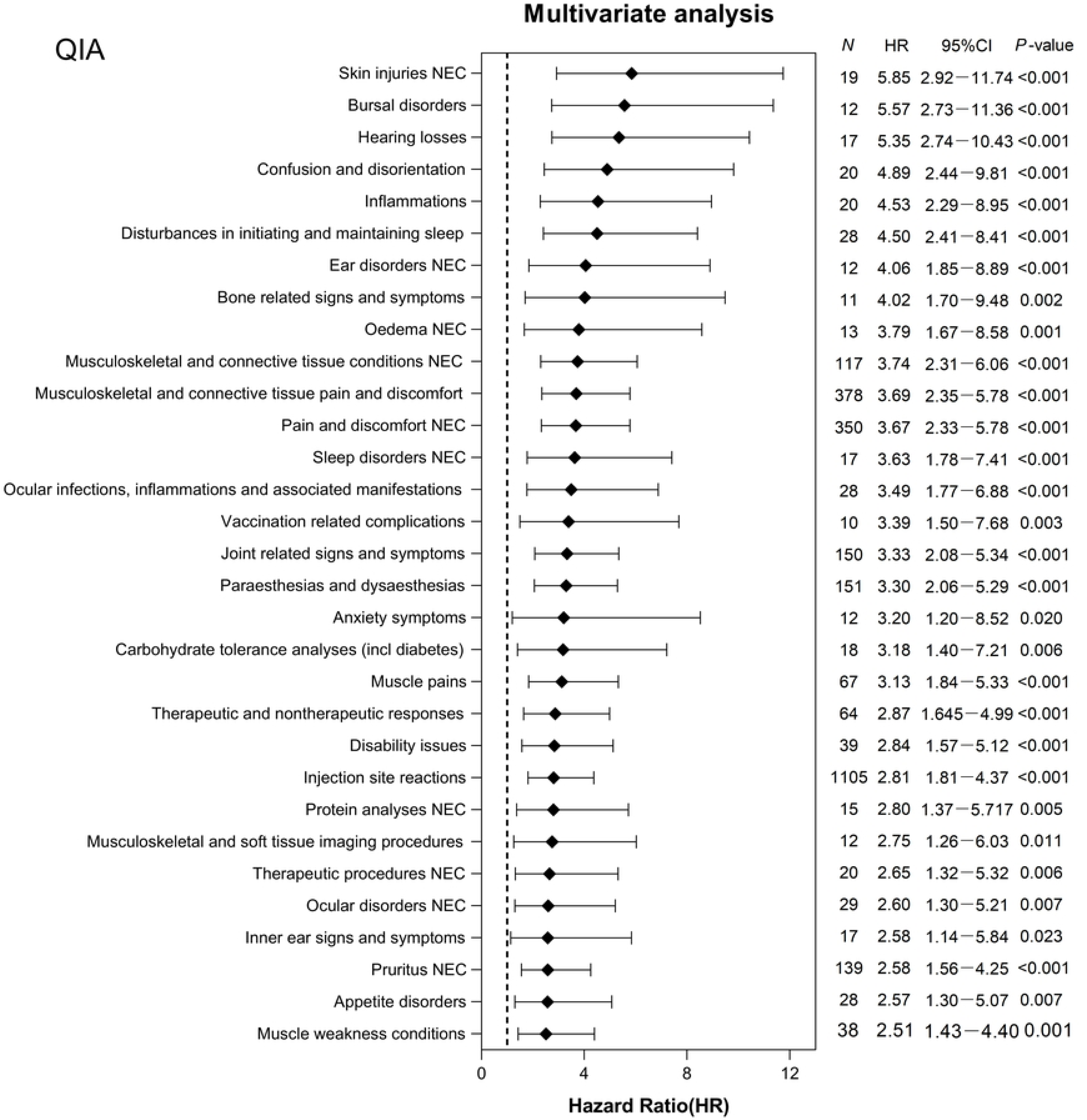
Adverse events (AEs) hazard ratios after quadrivalent influenza vaccines (QIA) vaccination estimated from a multivariate Cox proportional hazard model. The results showed that 57 quadrivalent influenza vaccine (QIA) vaccine-induced adverse events (AEs) with a statistically significant difference (*P*<0.05), estimated with a multivariate Cox proportional hazard regression analysis. N denotes the number of adverse cases in the non-recovered group; HR, hazard ratio; CI, confidence interval.

Four AEs were more common in the nonrecovery group: general disorders and administration site conditions (46.74%), musculoskeletal and connective tissue disorders (14.95%), skin and subcutaneous tissue disorders (12.17%), and nervous system disorders (9.63%). Among them, injection site reactions (20.00%; HR=2.81; 95% CI, 1.81–4.37), musculoskeletal and connective tissue pain and discomfort (6.84%; HR=3.69; 95% CI, 2.35–5.78), pain and discomfort NEC (6.34%; HR=3.67; 95% CI, 2.33–5.78), asthenic conditions (5.39%; HR=2.25; 95% CI, 1.41–3.58) typically exhibited a higher risk with QIA. Additionally, 28 AEs with FLUX were observed and five AEs were associated with the risk of developing AE, as estimated by the multivariate Cox regression model (Fig 3). The consistency index is 0.59. The adjusted HR distribution ranged from 2.83 to 6.31 and was subsequently categorized into three SOC terms (S6 Table). General disorders, administration site conditions (71.54%), and musculoskeletal and connective tissue disorders (23.58%) were the most frequent (similar to the TIA results). In particular, injection site reactions (67.48%; HR=2.83; 95% CI, 1.12–7.15), tissue condition NEC (13.01%; HR=3.64; 95% CI, 1.29–10.23), and joint-related signs and symptoms (10.57%; HR=3.75; 95% CI, 1.25– 11.26) were at heightened risk in the non-recovery group. A comparison of PSM before and after TIA, QIA, and FLUX vaccinations is shown in S2 Fig.

**Fig 3.**
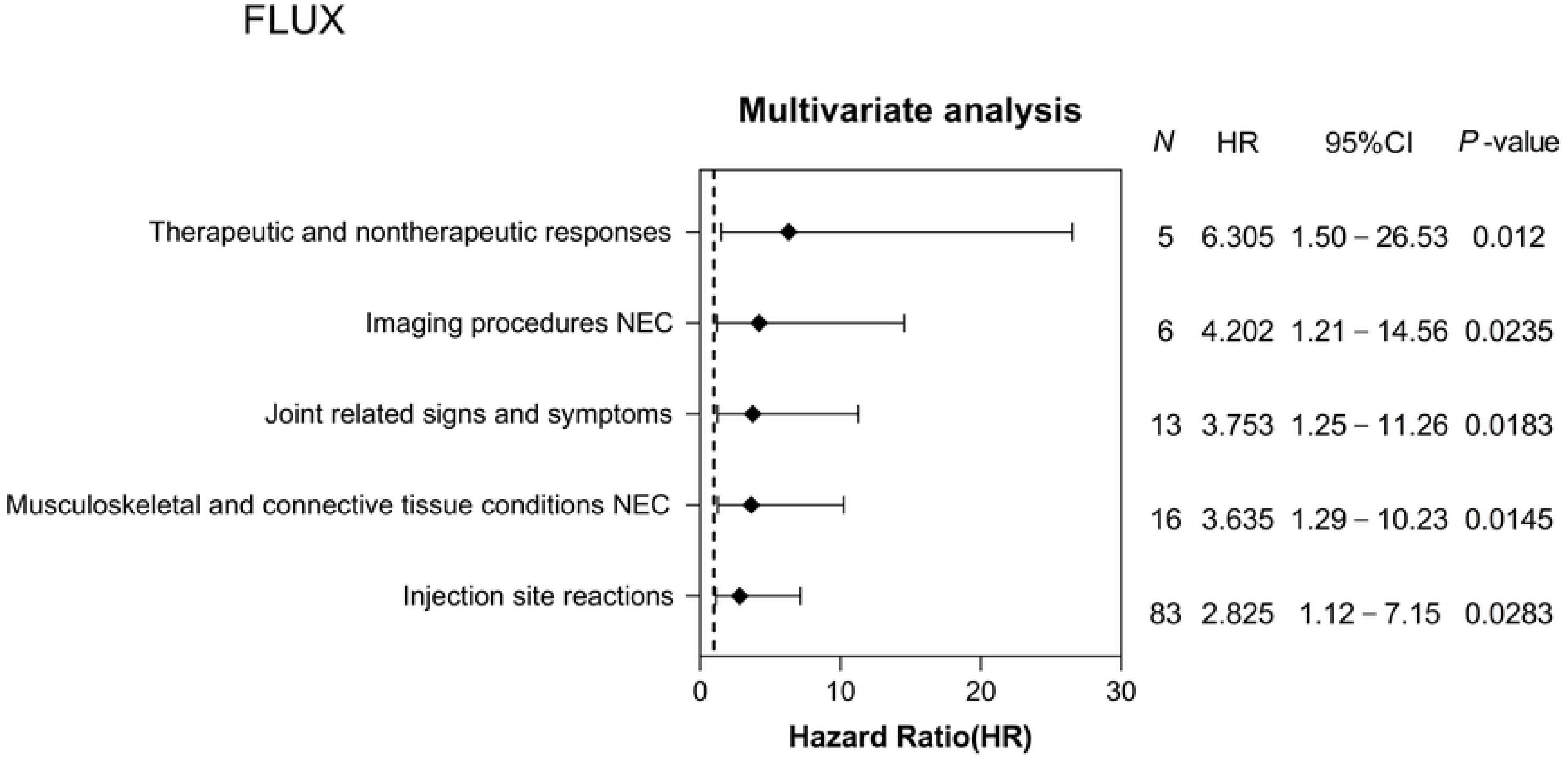
Adverse events (AEs) hazard ratios after influenza unknown manufacturer (FLUX) vaccination estimated from a multivariate Cox proportional hazard model. The results showed five influenza unknown manufacturer (FLUX) vaccine-induced adverse events (AEs) with a statistically significant difference (*P*<0.05), estimated using multivariate Cox proportional hazard regression analysis. N denotes the number of adverse cases in the non-recovered group; HR, hazard ratio; CI, confidence interval.

## Discussion

We analyzed whether major AEs influenced recovery following seasonal influenza vaccination in a hypertensive population. Among the 27 SOCs examined, four terms (general disorders and administration site conditions, musculoskeletal and connective tissue disorders, skin and subcutaneous tissue disorders, and nervous system disorders) were associated with a higher frequency of AEs in the PS-matched hypertensive population. Among the non-recovery group, the most common AEs were injection site reactions, musculoskeletal and connective tissue pain and discomfort, pain and discomfort NEC, and joint-related signs and symptoms. Most AEs (95.5%) occurred within 48 hours of vaccination. These results are consistent with those of several previous studies, suggesting that local and systemic reactions are common[32,33], and that all reported AEs were nonserious. Furthermore, our study showed results related to neurological aspects after seasonal influenza vaccination in the hypertensive population. Paraesthesias and dysesthesias (4.09%) were the main manifestations of TIA, and nervous system disorders were found in 9.63% with QIA; the most frequent AEs were headache NEC (3.33%), neurological signs and symptoms NEC (2.55%), paresthesia, and dysesthesia (2.73%). In addition, no association was found between FLUX and neurological aspects. Some studies have shown an association between the incidence of Greene-Barrow syndrome[33], Acute Disseminated Encephalomyelitis (ADEM), and seasonal influenza vaccination[34].

After TIA vaccination, there were 20 cases (0.88%) of disturbances in initiating and maintaining sleep, and 37 cases (1.63%) of NEC skin injuries in the population. Disturbances in initiating and maintaining sleep are psychiatric disorders; however, no studies have been conducted on seasonal influenza vaccines and psychiatric disorders. In a study evaluating the relationship between maternally inactivated influenza vaccination (IIV) and the risk of diagnosis of neurodevelopmental disorders in early childhood, no increased risk of neurodevelopmental disorders was found in children after maternal exposure to IIV[35]. The majority of rare AEs associated with QIA, including ear and labyrinth disorders (0.83%), eye disorders (1.03%), appetite disorders (0.51%), and respiratory, thoracic, and mediastinal disorders (0.83%), may hinder recovery in the hypertensive population. Rare AEs associated with vaccination with FLUX were primarily NEC (4.88%) in the hypertensive population.

Clinical trials are the gold standard to evaluate the safety and efficacy of new vaccines. However, they have limitations in evaluating rare AEs due to the short observation period and limited size of the testing populations. Post-marketing surveillance, such as a spontaneous reporting database, plays a critical role in continually monitoring the safety signals of AEs[36]. Nevertheless, our study had some limitations. First, the VAERS database suffers from underreporting, overreporting, missing data, and possible reporting based on indirect information such as media sources[32]. Thus, mild-to-moderate or severe AEs may be rarer than our findings[31]. Moreover, we cannot rule out the possibility of conditions or regulatory agency interventions due to unadjusted confounders, such as exposure to previous infections and individual occupational characteristics[37,38], which may affect the VAERS database information based on the year of reporting. Therefore, our findings may have involved unknown or invalidated AEs. Furthermore, the reduced sample size of the post-PSM study may have led to a selection bias, and we used a limited number of covariates that met the user-defined sample thresholds in the propensity score-matched target outcome.

## Conclusion

Our study provides insights into the AEs associated with seasonal influenza vaccination in hypertensive individuals, highlighting common local and systemic reactions. However, further research with larger and more diverse populations is needed to confirm these findings and better understand the impact of specific AEs on recovery in hypertensive populations. Additionally, ongoing post-marketing surveillance is essential to continuously monitor the safety signals of AEs and to inform future vaccination strategies.

## Data Availability

The raw data used in this study are available at https://vaers.hhs.gov/data/datasets.html.

## Supporting information

## S1 Appendix.

(DOCX)

## Acknowledgments

The authors thank the study participants involved in this study.

## Author contributions

**Conceptualization:** Hao Wu, Xiaona He

**Data curation:** Hao Wu

**Formal analysis:** Hao Wu, Yu Cao

**Methodology:** Hao Wu

**Funding acquisition:** Wei Gao

**Supervision:** Wei Gao

**Project administration:** Wei Gao

**Writing-original draft:** Hao Wu

**Writing-review & editing:** Wei Gao

## References

[1] World Health Organization. Influenza (Seasonal). https://www.who.int/news-room/fact-sheets/detail/influenza-(seasonal) (accessed 29 August 2024).

[2] Hutchinson EC. Influenza Virus. Trends Microbiol. 2018;26:809–10. 10.1016/J.TIM.2018.05.013.

[3] Martini M, Gazzaniga V, Bragazzi NL, Barberis I. The Spanish Influenza Pandemic: a lesson from history 100 years after 1918. J Prev Med Hyg. 2019;60:E64–7. 10.15167/2421-4248/JPMH2019.60.1.1205.

[4] Akin L, Gözel MG. Understanding dynamics of pandemics. Turk J Med Sci. 2020;50:515–9. 10.3906/SAG-2004-133.

[5] Li J, Zhang Y, Zhang X, Liu L. Influenza and Universal Vaccine Research in China. Viruses. 2022;15. 10.3390/V15010116.

[6] Czaja CA, Miller L, Alden N, Wald HL, Cummings CN, Rolfes MA, et al. Age-Related Differences in Hospitalization Rates, Clinical Presentation, and Outcomes Among Older Adults Hospitalized With Influenza-U.S. Influenza Hospitalization Surveillance Network (FluSurv-NET). Open Forum Infect Dis. 2019;6. 10.1093/OFID/OFZ225.

[7] Holstein R, Dawood FS, O’Halloran A, Cummings C, Ujamaa D, Kirley PD, et al. Characteristics and Outcomes of Hospitalized Pregnant Women With Influenza, 2010 to 2019 : A Repeated Cross-Sectional Study. Ann Intern Med. 2022;175:149–58. 10.7326/M21-3668.

[8] Olson SM, Newhams MM, Halasa NB, Feldstein LR, Novak T, Weiss SL, et al. Vaccine Effectiveness Against Life-Threatening Influenza Illness in US Children. Clinical Infectious Diseases. 2022;75:230–8. 10.1093/CID/CIAB931.

[9] Andrejko KL, Myers JF, Openshaw J, Fukui N, Li S, Watt JP, et al. Receipt of COVID-19 and seasonal influenza vaccines in California (USA) during the 2021-2022 influenza season. Vaccine. 2023;41:1190–7. 10.1016/J.VACCINE.2022.12.052.

[10] Harding AT, Heaton NS. Efforts to Improve the Seasonal Influenza Vaccine. Vaccines (Basel). 2018;6. 10.3390/VACCINES6020019.

[11] McKeage K. Inactivated quadrivalent split-virus seasonal influenza vaccine (Fluarix® Quadrivalent): A review of its use in the prevention of disease caused by influenza A and B. Drugs. 2013;73:1587–94. 10.1007/s40265-013-0114-3.

[12] Fens T, De Boer PT, Van Puijenbroek EP, Postma MJ. Inclusion of Safety-Related Issues in Economic Evaluations for Seasonal Influenza Vaccines: A Systematic Review. Vaccines (Basel). 2021;9:1–28. 10.3390/VACCINES9020111.

[13] Asturias EJ, Wharton M, Pless R, MacDonald NE, Chen RT, Andrews N, et al. Contributions and challenges for worldwide vaccine safety: The Global Advisory Committee on Vaccine Safety at 15 years. Vaccine. 2016;34:3342–9. 10.1016/J.VACCINE.2016.05.018.

[14] Shimabukuro TT, Nguyen M, Martin D, DeStefano F. Safety monitoring in the Vaccine Adverse Event Reporting System (VAERS). Vaccine. 2015;33:4398–405. 10.1016/J.VACCINE.2015.07.035.

[15] Varricchio F, Iskander J, Destefano F, Ball R, Pless R, Braun MM, et al. Understanding vaccine safety information from the Vaccine Adverse Event Reporting System. Pediatr Infect Dis J. 2004;23:287–94. 10.1097/00006454-200404000-00002.

[16] World Health Organization. Causality assessment of an adverse event following immunization (AEFI): user manual for the revised WHO classification, 2nd ed., 2019 update. https://www.who.int/publications/i/item/9789241516990 (accessed 29 August 2024).

[17] World Health Organizatio. Global manual on surveillance of adverse events following immunization. https://www.who.int/publications/i/item/9789241507769 (accessed 29 August 2024).

[18] Newall AT, Chaiyakunapruk N, Lambach P, Hutubessy RCW. WHO guide on the economic evaluation of influenza vaccination. Influenza Other Respir Viruses. 2018;12:211–9. 10.1111/IRV.12510.

[19] Chen Q, Wang L, Xie M, Li X. Recommendations for influenza and Streptococcus pneumoniae vaccination in elderly people in China. Aging Med (Milton). 2020;3:1–11. 10.1002/AGM2.12102.

[20] Clar C, Oseni Z, Flowers N, Keshtkar-Jahromi M, Rees K. Influenza vaccines for preventing cardiovascular disease. Cochrane Database Syst Rev. 2015;2015. 10.1002/14651858.CD005050.PUB3.

[21] Recommended adult immunization schedule for ages 19 years or older: United States, 2022. JAAPA. 2022;35:1–22. 10.1097/01.JAA.0000822520.39079.BD.

[22] Feng L, Peng Z, Wang D, Yang P, Yang J, Zhang Y, et al. Technical guidelines for seasonal influenza vaccination in China, 2018-2019. Zhonghua Liu Xing Bing Xue Za Zhi. 2018;39:1413–25. 10.3760/CMA.J.ISSN.0254-6450.2018.11.001.

[23] Centers for Disease Control and Prevention. National Center for Immunization and Respiratory Diseases (NCIRD). https://www.cdc.gov/about/leadership/leaders/ncird.html (accessed 29 August 2024).

[24] Li Q, Zhang M, Chen H, Wu F, Xian J, Zheng L, et al. Influenza Vaccination Coverage among Older Adults with Hypertension in Shenzhen, China: A Cross-Sectional Survey during the COVID-19 Pandemic. Vaccines (Basel). 2021;9. 10.3390/VACCINES9101105.

[25] Wang Q, Yue N, Zheng M, Wang D, Duan C, Yu X, et al. Influenza vaccination coverage of population and the factors influencing influenza vaccination in mainland China: A meta-analysis. Vaccine. 2018;36:7262–9. 10.1016/J.VACCINE.2018.10.045.

[26] Centers for Disease Control and Prevention. Seasonal Influenza Vaccine Safety: A Summary for Clinicians. https://www.cdc.gov/flu/professionals/vaccination/vaccine_safety.htm (accessed 29 August 2024).

[27] S. MS, V. SM, M. TN, M. Š-ZM. Medical dictionary MedDRA: Used in over 60 countries, among which is Montenegro. Hospital Pharmacology. 2015;2:266–71. 10.5937/HPIMJ1502266M.

[28] MedDRA. What’s New with MedDRA Version 26.1. https://admin.meddra.org/sites/default/files/guidance/file/whatsnew_26_1_English.pdf (accessed 29 August 2024).

[29] Brown EG, Wood L, Wood S. The medical dictionary for regulatory activities (MedDRA). Drug Saf. 1999;20:109–17. 10.2165/00002018-199920020-00002.

[30] So CS, Jin SH, Kang WS. Propensity-Score-Matched Evaluation of Adverse Events Affecting Recovery after COVID-19 Vaccination: On Adenovirus and mRNA Vaccines. Vaccines (Basel). 2022;10. 10.3390/VACCINES10020284.

[31] Ali MS, Prieto-Alhambra D, Lopes LC, Ramos D, Bispo N, Ichihara MY, et al. Propensity score methods in health technology assessment: Principles, extended applications, and recent advances. Front Pharmacol. 2019;10. 10.3389/FPHAR.2019.00973/PDF.

[32] Tebaa A, Benkirane R, Alj L, Cherkaoui I, Soulaymani-Bencheikh R. Monitoring the safety of influenza A/H1N1 pandemic and seasonal vaccines in Morocco. Ther Adv Vaccines Immunother. 2022;10. 10.1177/25151355221088157.

[33] Gattás VL, Braga PE, Koike ME, Lucchesi MBB, de Oliveira MMM, Piorelli R de O, et al. Safety assessment of seasonal trivalent influenza vaccine produced by Instituto Butantan from 2013 to 2017. Rev Inst Med Trop Sao Paulo. 2018;61. 10.1590/S1678-9946201961004.

[34] Fujimori M, Nakamura M. Association between seasonal influenza vaccines and the increased risk of acute disseminated encephalomyelitis, estimated using the Vaccine Adverse Event Reporting System. Pharmazie. 2022;77:262–9. 10.1691/PH.2022.2354.

[35] Foo D, Sarna M, Pereira G, Moore HC, Regan AK. Association between maternal influenza vaccination and neurodevelopmental disorders in childhood: a longitudinal, population-based linked cohort study. Arch Dis Child. 2023;108:647–53. 10.1136/ARCHDISCHILD-2022-324269.

[36] Du J, Xiang Y, Sankaranarayanapillai M, Zhang M, Wang J, Si Y, et al. Extracting postmarketing adverse events from safety reports in the vaccine adverse event reporting system (VAERS) using deep learning. Journal of the American Medical Informatics Association. 2021;28:1393–400. 10.1093/JAMIA/OCAB014.

[37] Thompson MG, Stenehjem E, Grannis S, Ball SW, Naleway AL, Ong TC, et al. Effectiveness of Covid-19 Vaccines in Ambulatory and Inpatient Care Settings. New England Journal of Medicine 2021;385:1355–71. 10.1056/NEJMOA2110362/SUPPL_FILE/NEJMOA2110362_DATA-SHARING.PDF.

[38] Mosterín Höpping A, McElhaney J, Fonville JM, Powers DC, Beyer WEP, Smith DJ. The confounded effects of age and exposure history in response to influenza vaccination. Vaccine. 2016;34:540–6.10.1016/J.VACCINE.2015.11.058.

